# Domestic and international mobility trends in the United Kingdom during the COVID-19 pandemic: An analysis of Facebook data

**DOI:** 10.1101/2021.08.03.21261545

**Authors:** Harry ER Shepherd, Florence S Atherden, Ho Man Theophilus Chan, Alexandra Loveridge, Andrew J Tatem

## Abstract

Since early March 2020, the COVID-19 epidemic across the United Kingdom has led to a range of social distancing policies, which resulted in changes to mobility across different regions. An understanding of how these policies impacted travel patterns over time and at different spatial scales is important for designing effective strategies, future pandemic planning and in providing broader insights on the population geography of the country. Crowd level data on mobile phone usage can be used as a proxy for population mobility patterns and provide a way of quantifying in near-real time the impact of social distancing measures on changes in mobility. Here we explore patterns of change in densities, domestic and international flows and co-location of Facebook users in the UK from March 2020 to March 2021. We find substantial heterogeneities across time and region, with large changes observed compared to pre-pandemic patterns. The impacts of periods of lockdown on distances travelled and flow volumes are evident, with each showing variations, but some significant reductions in co-location rates. Clear differences in multiple metrics of mobility are seen in central London compared to the rest of the UK, with each of Scotland, Wales and Northern Ireland showing significant deviations from England at times. Moreover, the impacts of rapid changes in rules on international travel to and from the UK are seen in substantial fluctuations in traveller volumes by destination. While questions remain about the representativeness of the Facebook data, previous studies have shown strong correspondence with census-based data and alternative mobility measures, suggesting that findings here are valuable for guiding strategies.

## Introduction

The COVID-19 pandemic and measures to mitigate it have resulted in unprecedented changes to population dynamics around the World (Banholzer et al., 2021; Flaxman et al., 2020; Hsiang et al., 2020; Lai et al., 2020; Ruktanonchai et al., 2020; Unwin et al., 2020). Most countries have implemented prevention policies such as regional lockdowns and social distancing (Kissler et al., 2020; Thakkar et al., 2020; Wellenius et al., 2020) to slow the spread of the disease and limit impacts on health systems. The impact of these policies on population dynamics is unclear and likely varies significantly by geography, demographics and over time (Buckee et al., 2020; Buckee et al., 2021; Gibbs et al., 2021). Understanding these impacts and relationships is important for guiding ongoing efforts to mitigate the effects of the COVID-19 pandemic, as well as for future pandemic preparedness planning.

Over the 12 months from March 2020, the UK recorded more than 4 million cases of COVID-19, resulting in around 120,000 deaths (Public Health England, 2021). A wide variety of national and regional restrictions were put in place throughout the course of 2020 and into early 2021. This involved national stay at home ‘lockdown’ measures in March 2020, November 2020 and January 2021. Additionally, England, Scotland, Wales and Northern Ireland all implemented differing restrictions at different times, with regionally stratified ‘tiered’ systems. Moreover, international travel restrictions in and out of the UK were under constant review and change, as situations changed domestically and abroad. Many of these mitigation policies involved restrictions relating to social contact and travel, but the effects of these on changes in population densities, travel patterns and mixing at fine spatial and temporal scales have not been examined together across the course of the pandemic.

Multiple sources of digital data on proxies for population mobility and behaviours have been put forward and used to examine changing dynamics and understand the impacts of disease mitigation measures (Gonzalez et al., 2008; Jeffrey et al., 2020). During the course of the COVID-19 pandemic, many new sources of mobility data have been utilised and made available, principally deriving from mobile phones and their usage. These include mobility data and reports made available by technology giants such as Google (Google, 2020), Apple (Apple, 2020) and Facebook (Facebook, 2020; Maas, 2019). Many studies have examined social connectedness (Chang et al., 2021; Fritz & Kauermann, 2020; Kuchler et al., 2020; Lai et al., 2020) or human movement (Beria & Lunkar, 2021; Ruktanonchai et al., 2020; Galeazzi et al., 2020; Gibbs et al., 2021; Kraemer et al., 2021) and their relation to the dynamics and spread of COVID-19.

Here we explore aggregated and anonymised data from Facebook on the movement patterns of active users in the UK using geolocation services during the period March 2020 - March 2021. These data are made available at fine temporal and spatial scales, in near-real time and in multiple forms, enabling insights not possible with many other sources of mobility data. We examine the changes in domestic movement patterns and densities compared to a pre-pandemic baseline, co-location probabilities, as well as variations in international travel in and out of the UK.

## Data and methods

### Facebook data

Facebook disease prevention maps (Facebook, 2021) provide information on the location, movement and interactions of active Facebook users. The data is produced using geolocation (GPS) services from mobile devices with the Facebook app installed and with location services switched on. The location of Facebook users is mapped using a Bing tile architecture (Microsoft, 2018), assigning users to individual tiles. The minimum tile size available (level 16) is approximately 600m x 600m. However, due to the constraints in collating location data from a large number of Facebook users, mobility data is commonly supplied by Facebook at lower resolutions (greater tile sizes). For datasets that use administrative regions, tile data is then aggregated by Facebook using Pitney-Bowes polygon boundaries (Lyer et al., 2020; Precisely, 2020).

In this paper we include data from four available datasets: population density (tile level), movement between tiles, co-location and international travel, spanning the period from 10th March 2020 to 9th March 2021. This encompasses three periods of tight restrictions that included stay-at-home orders and closure of many businesses by the UK government in England, which we here term ‘lockdowns’: 23rd March 2020 - 12th May 2020; 5th November 2020 - 1st December 2020; and 6th January 2021 - 9th March 2021. These are from now on referred to as lockdown one, lockdown two and lockdown three, respectively. Similar restrictions were imposed at the same time (or at very similar times) in Wales, Scotland and Northern Ireland.

Facebook provides the data in a range of time periods. Population density and movement between tiles datasets are provided daily across three 8-hour time periods: 00:00 - 08:00, 08:00 - 16:00 and 16:00 - 00:00. These will be referred to as nighttime, daytime and evening respectively. International travel data is provided daily and the co-location data is provided weekly. All time periods are kept as Coordinated Universal Time (UTC). Therefore between 29th March 2020 and 24th October 2020, the 8-hour time periods represent 01:00 - 09:00, 09:00 - 17:00 and 17:00 - 01:00 in the UK due to the shift from Greenwich mean time (GMT) into British summer time (BST). In addition to this data, Facebook also provides baseline values of mobility for certain datasets. They are calculated prior to the introduction of government measures on population mobility in order to allow comparisons between pre-COVID and current movements of Facebook users.

There are a number of measures applied to the data to ensure the privacy and anonymity of Facebook users that may also impact the interpretation of the mobility data. Firstly, only Facebook users with location services actively enabled within the specified time period are included in each dataset. In addition, thresholds for the number of individuals recorded in each dataset are applied. In the population density, movement between tile and co-locations datasets, this is 10 users per tile, movement vector or administrative region respectively. In the international movement dataset, there is a minimum threshold of 1000 users moving between two countries per day. Finally, absolute values of Facebook user mobility are not able to be shared publicly. Instead, values of relative change are calculated using the data, often comparing baseline values to current mobility of Facebook users. Together, this ensures the identity and security of Facebook users is maintained.

Combining Facebook data into larger aggregations can provide human mobility data relevant to pre-established administrative boundaries. In this paper we aggregate Facebook data using two classified systems: Nomenclature of territorial units for statistics (NUTS) level 1 regions (Figure 1) (December 2020 boundaries; Office for National Statistics, 2017) and UK Local Authority Districts (LAD) (April 2020 boundaries; Office for National Statistics, 2020). When aggregating data, we took the centroid coordinate of each Bing tile and matched it with the LAD/NUTS region in which it fell. We then summed the data within each aggregation. It is important to note that aggregating data does not overcome disclosure issues as these are applied at tile level by Facebook before sharing with external partners. Additionally, due to the mismatch between the shape of Bing tiles and the boundaries of the aggregations, a marginal error is expected when using this method that is unavoidable given the structure of the data. To account for some of the mismatch caused when aggregating tiles to LAD, we used a population weighted method of assigning tiles to large population centres. We did this on occasions when, due to mismatches between tiles and LAD boundaries, areas with expected high population densities (e.g. London LADs) had no Bing tiles assigned to them. Tile to aggregation lookups are provided in the supplementary materials.

**Figure 1.**
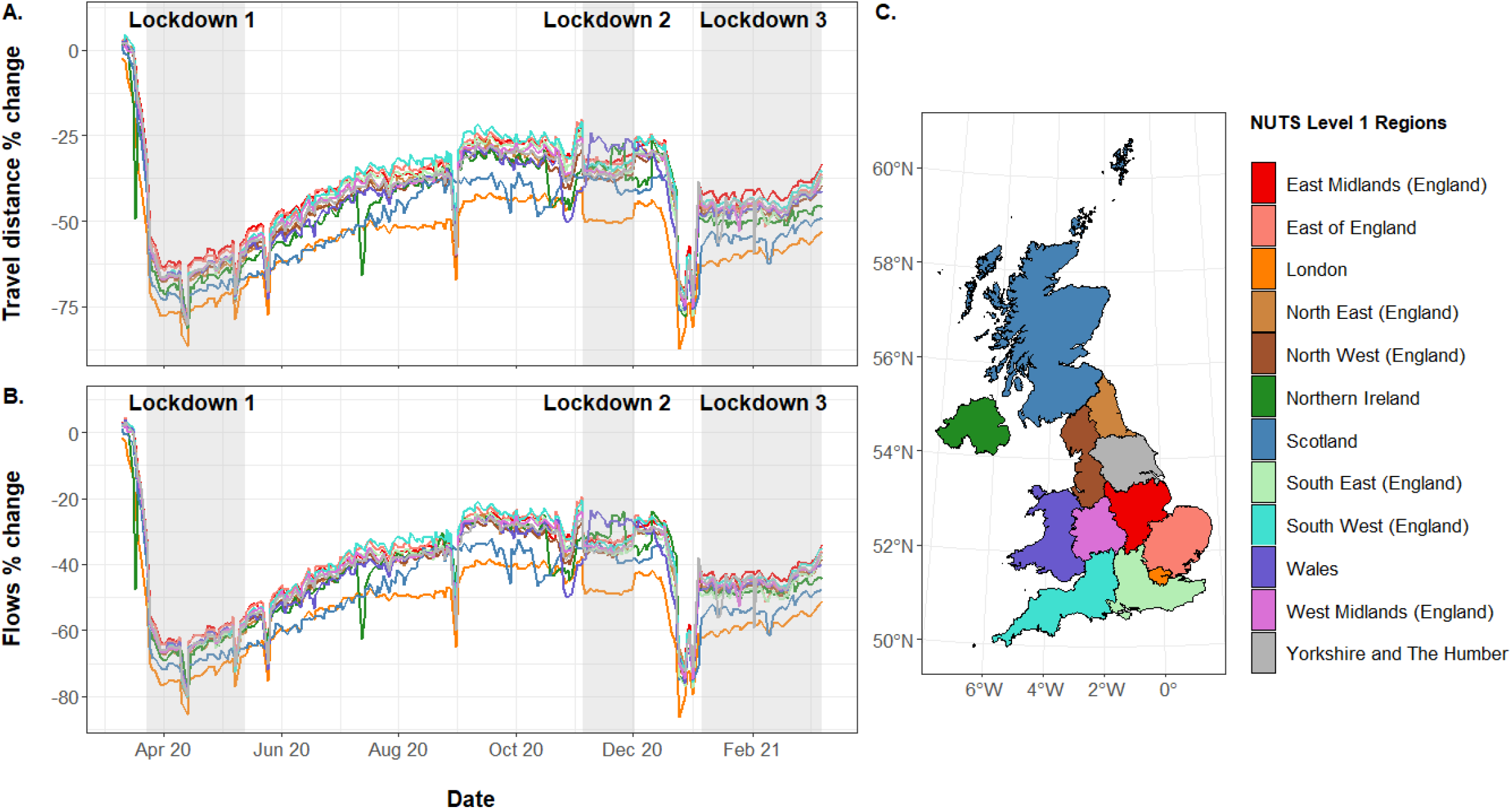
Percentage changes in (A) distances travelled and (B) population movement (flows) of Facebook users within (C) UK NUTS level 1 regions. Journeys included are those made that begin during 00:00 - 08:00 and end during 08:00 - 16:00 on weekdays between March 10th 2020 and March 9th 2021. See supplementary material for other time periods and weekends. The change is relative to a pre-COVID baseline (see Methods for details). Regions follow NUTS level 1 ultra generalised clipped boundaries in the UK (Office for National Statistics, 2019).

The data used in this study are proprietary data owned by Facebook. The data are not publicly available but are available to researchers by application to the Facebook Data for Good Partner Program (https://dataforgood.fb.com/). The research was approved by the University of Southampton Ethics and Research Governance committee (Ref.48113).

### Changes in densities

The population density (tile level) dataset represents the average number of Facebook users within a given Bing tile over an 8-hour time period. The tile resolution of the population density data provided varies between datasets. For Britain-wide population density, Bing tile level 13 data is provided. For smaller regions (e.g. individual cities such as London), data can be provided by Facebook down to Bing tile level 16.

For each day, population density data is provided in the three 8-hour time periods. Within each time period, active Facebook users are assigned to their modal Bing tile. Tiles that do not reach 10 users are removed from the dataset. For each tile that overcomes the user threshold, data is provided on the tile location and the average number of active Facebook users assigned to that tile. Additionally, for each tile within each day of the week and individual time period that is included, a single baseline value is provided. This is calculated from the average population density of Facebook users over a 90-day period up to and including the 9th March 2020. As there are 7 weekdays and 3 time periods per day, each tile has a total of 21 baseline values.

In order to determine the impact of mobility measures on the population density of Facebook users across the UK we first extracted tile level 13 data for the entirety of the UK. This data was then aggregated to NUTS level 1 to compare the impact of the measures across different UK regions. Then, to examine how these changes were reflected at smaller spatial scales, we extracted tile level 16 data for the entirety of London. For both regional and London analysis of Facebook user population density we calculated values of change relative to the pre-COVID baseline period.

### Changes in domestic mobility

The UK movement between tiles (flows) data represents the movement of Facebook users between two UK Bing tiles across two adjacent 8-hour time periods. For each 8-hour time period, the Bing tile where a Facebook user spends most of their time (modal tile) is recorded. This is repeated at the following 8-hour time period to produce a vector between two Bing tiles. The number of Facebook users that produce the same vector between two time periods is summed to give a total number of users that moved between two Bing tiles across the same two 8-hour time periods. The Euclidean distance between these tiles is also provided. If less than 10 Facebook users are recorded as travelling across an individual vector the data is withheld by Facebook. In addition to the number of people moving between two tiles, a baseline value is provided for each tile to tile vector that crosses the privacy threshold. This is the average number of Facebook users that moved between two tiles in the same two adjacent time periods per weekday in the 45 day period up to and including 9th March 2020.

In order to examine changes in the movement of Facebook users within the UK, we extracted tile level 12 data for movement of Facebook users. We aggregated this data to LADs and separated journeys into those moving between two different tiles in the same LADs (within LAD) and between two different LADs (between LAD). In addition, we aggregated the number of Facebook users who were present in the same modal tile between two adjacent time periods. Maintaining the two LAD journey types, we then calculated the total distance travelled by Facebook users by multiplying each tile to tile vector volume by the length between the two tiles. Each distance was then summed to give a total distance travelled between or within LADs. We then calculated the relative change in flows/distance travelled relative to the baseline number of Facebook users.

### Co-location

Facebook co-location is the probability that two Facebook users from different Pitney-Bowes administrative regions are present within the same level 16 Bing tile for a random minute during one week. To calculate this metric, Facebook assigns users to the administrative unit where they are consistently observed overnight over the course of one week (i.e. home location). Users that do not have a consistent overnight location are removed from the dataset. Facebook then calculates the probability that a user from two different home locations are present within the same level 16 Bing tile within a 5-minute time bin during one-week. Facebook does not indicate where or precisely when co-location events occur. For example, the co-location rate for Southampton and London sums over co-location events that happen in Southampton, in London and in all other administrative units to give a single value of co-location between Southampton and London.

In order to calculate the co-location probability of Facebook users within different UK regions, we matched administrative regions to their NUTS 1 region (Figure 1c). For each NUTS 1 region, we calculated the weekly mean of the log-value of all co-location probabilities where at least one of the Facebook users home locations was within that region. For example, the weekly mean log co-location value of Northern Ireland is the average log co-location value of all weekly co-location probabilities where at least one user’s home location was within an administrative boundary in Northern Ireland. For each NUTS level 1 region, this provides the probability of a Facebook user who’s home location is within that region spends one-minute within a level 16 Bing tile with a Facebook user from a different administrative boundary.

### Changes in international travel

International travel pattern data consists of the daily number of Facebook users moving between different countries. To produce this, Facebook calculates the number of unique Facebook users with location services enabled travelling from the origin to the destination country each day (date defined by UTC time zone). For data to be provided, a minimum of 1000 unique Facebook users is required for each country pair.

To determine weekly changes to international travel, we extracted all journeys originating from and ending in the UK. We then determined the ten countries with the highest travel volumes for travel to and from the UK respectively. Values of change in travel volumes were calculated relative to the highest mean weekly movement observed across the study period. This was calculated separately for travel in and out of the UK (Highest mean weekly movement out: UK - Ireland, week commencing 10th August 2020; Highest mean weekly movement in: Spain - UK, week commencing 10th March 2020).

## Results

Analyses show substantial changes in mobility patterns over the course of the March 2020 - March 2021 period, with strong regional and international differences. Here we present summary findings from the analysis of Facebook user data, with additional outputs and analyses presented in supplemental materials.

Figure 1 shows percentage changes in flows and distances travelled by Facebook users over the study period, broken down by NUTS level 1 regions for journeys ending between the 08:00 - 16:00 period. It highlights some of the substantial impacts that the pandemic and mitigation restrictions had on mobility across the country, with substantial changes seen relative to the baseline period (the 90 days up to and including 9th March 2020). Figure 1 suggests that the first national lockdown had the largest impact on population mobility, with both the percentage change in flows and distances travelled decreasing by about 75%. While the second and third lockdown had minimal impact on the percentage changes in flows and distances travelled, it slightly reduced the fluctuation in mobility patterns. The second lockdown appears to have had the smallest impact on population mobility as mobility during the period remained similar to the levels just prior to it. Little evidence is seen in Figure 1 of changes to mobility patterns due to the regional restrictions put in place in the East Midlands and North West in England over the 2-3 months prior to the second lockdown, with both of those regions showing very similar patterns to those areas not put under such restrictions.

The patterns of percentage change in distances travelled and percentage change in flows for weekdays (Monday to Friday) during the daytime (08:00 - 16:00) are almost identical. Over the period of the first lockdown, as indicated by the shaded region, a substantial drop in percentage changes for both flows and travel distance is seen. As restrictions for the first lockdown ease, a gradual increase is observed for both percentage changes in flows and distances travelled, with fluctuations in between. Unlike the first lockdown, there was not a significant decrease in percentage change in flows and distances travelled across the second lockdown. It is apparent that there was little change in distances travelled and population movement. In late December 2020, there was a sudden drop in percentage change in flows and distances travelled prior to the third lockdown; this may reflect the differing travel patterns over the Christmas holiday period, with a percentage drop of 87% on 25th December and above 75% for most of the 24th - 29th December period. Over the third lockdown, population movement was stable, similar to the second lockdown.

Figure 1 also shows that administrative units within the London region typically show the largest deviation from general patterns. While most regions have higher or similar flows and distance travelled to the baseline period (i.e. closer to a percentage change of zero), the London region has a much lower percentage change for most of the study period. As well as this, across the study period, Wales displays patterns in movement and travel similar to the other regions. However, over the second lockdown, Wales displays the greatest percentage change for both flows and distances travelled. Finally, Scotland does not follow the occasional dips in percentage change in flows and distances travelled with the other regions.

Panels A and B in Figure 2 show that both percentage changes in flows and distance travelled within local authority districts (LAD) and between LAD over weekdays are very similar throughout the study period. Very much like Figure 1, the largest decrease in percentage changes is observed over the first lockdown. Small changes over the second lockdown are seen and there were large decreases prior to the third lockdown. Over the second and third lockdown period, both percentage changes in flows and distances travelled were generally constant. Journeys that were between LAD showed slightly lower percentage changes compared to journeys within LAD.

**Figure 2.**
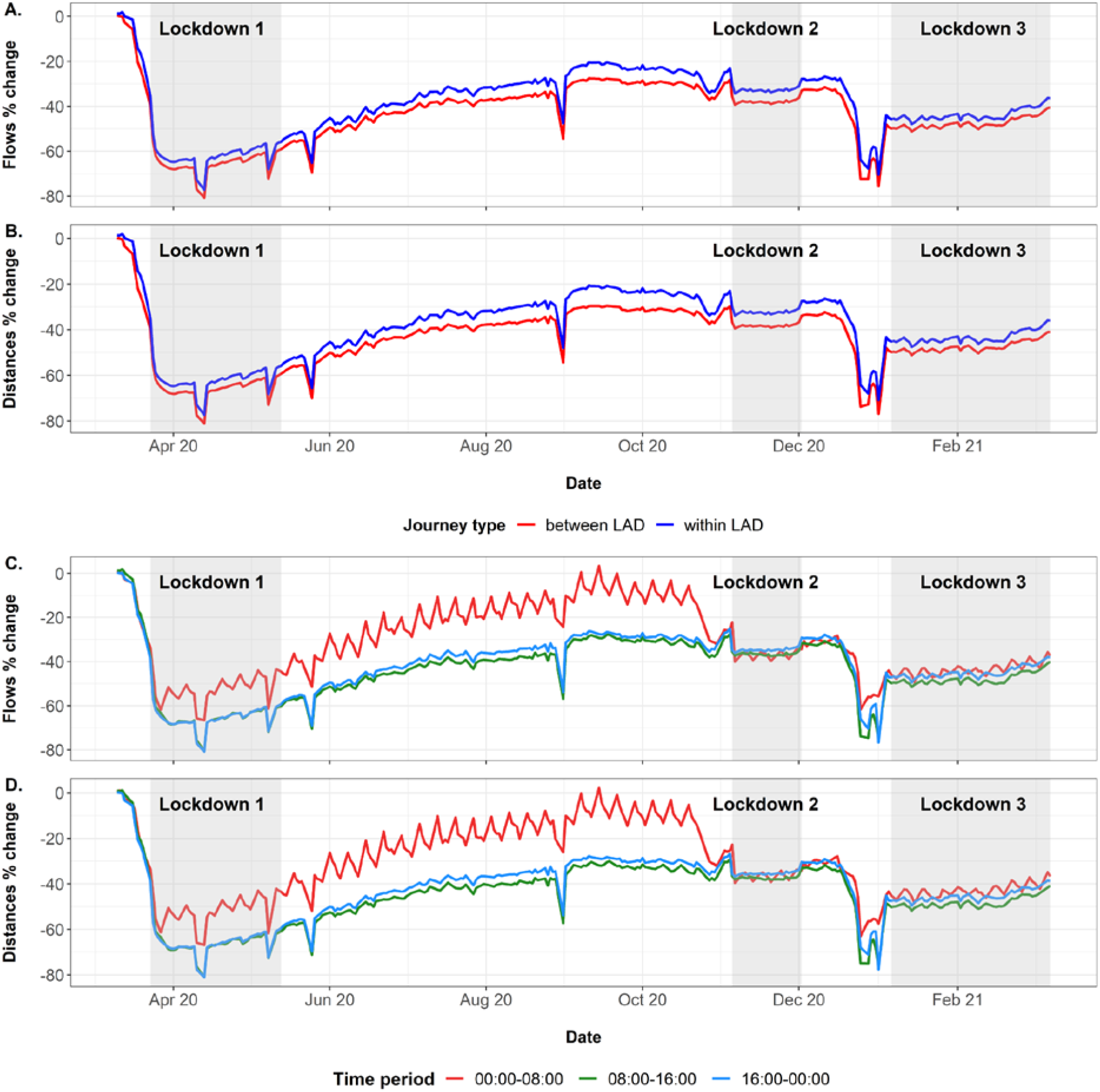
Percentage changes in (A, C) daily movement (flows) of Facebook users and (B, D) total distance travelled on weekdays between March 10th 2020 and March 9th 2021. A and B show the changes corresponding to different journey types (between and within UK local authority districts). C and D show the changes corresponding to all journeys across adjacent 8-hour time periods. Time period shown represents the second time period, where the journey ends. The change is relative to a pre-COVID baseline (see Methods for details). See supplementary material for changes to weekend flows and distances.

Panels C and D of Figure 2 show how changes in flows and distances vary by period of the day. Changes in both human mobility and distances travelled for the GMT time periods 08:00 - 16:00 and 16:00 - 00:00 showed similar patterns with 08:00 - 16:00 having a slightly lower percentage change in both flows and distances travelled. The 00:00 - 08:00 period shows some strong variations, which converge with the patterns of the other two time periods for the rest of the study period after the second lockdown period. The figure is impacted strongly by the shift to British Summer Time (BST) between 29th March 2020 and 24th October 2020, where the 8-hour time periods changed to represent 01:00 - 09:00, 09:00 - 17:00 and 17:00 - 01:00. This meant that the 00:00 - 08:00 GMT period switched to represent 01:00 - 09:00 BST, and therefore captured a greater amount of commuting/school-related travel, diverging from the other two periods during BST, when compared against GMT baseline periods. Nevertheless, the fluctuations for weekends and national holidays remain apparent throughout, with stronger impacts during the nighttime period apparent.

Figure 3 examines trends in co-location probabilities by NUTS level 1 regions over the study period. The general pattern here shows similarities to Figures 1 and 2, but also with notable differences. Unsurprisingly, co-location probabilities were substantially lower throughout most of the study period than those in early March prior to restrictions. Each lockdown was associated with drops in probabilities, though generally the drops for lockdowns two and three were not as substantial or low. However, the impact of the lockdown restrictions, especially for periods two and three, are more evident in co-location than mobility measures. Except for early March 2020, the summer and Christmas holiday periods consistently had the highest probabilities.

**Figure 3.**
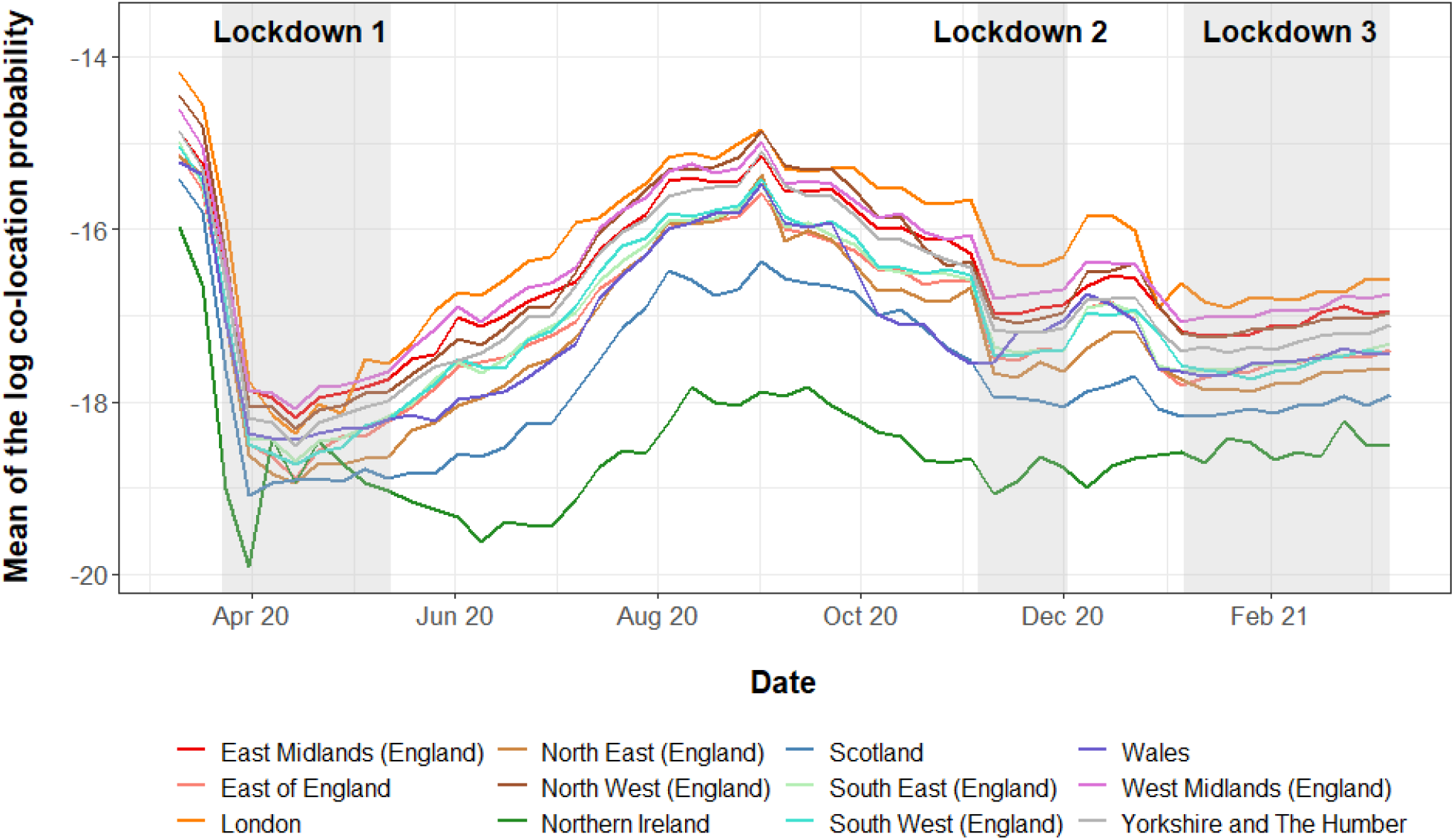
Weekly average log co-location probabilities for Facebook users whose home location is within different UK NUTS level 1 regions. Data included between March 10th 2020 and March 9th 2021.

While for most of the observed dates, all the NUTS level 1 regions of the UK exhibit similar co-location trends, Northern Ireland showed the lowest probabilities compared to the other regions, whereas the region consistently highest was London. Similar to the observations from Figures 1 and 2 above, during the lockdown periods changes tended to be minimal, but with gradual increases seen.

Figures 1 - 3 highlight changes that occurred over time and regionally in relation to the mobility and co-locations of Facebook users. In particular, the London region was highlighted as an outlier and Figures 1 and 2 emphasise the switch that occurred across the country in terms of increasing local movements over long distance travel. Figure 4 shows patterns of Facebook user densities at fine spatial scales for London and its surrounding area for four time periods.

**Figure 4.**
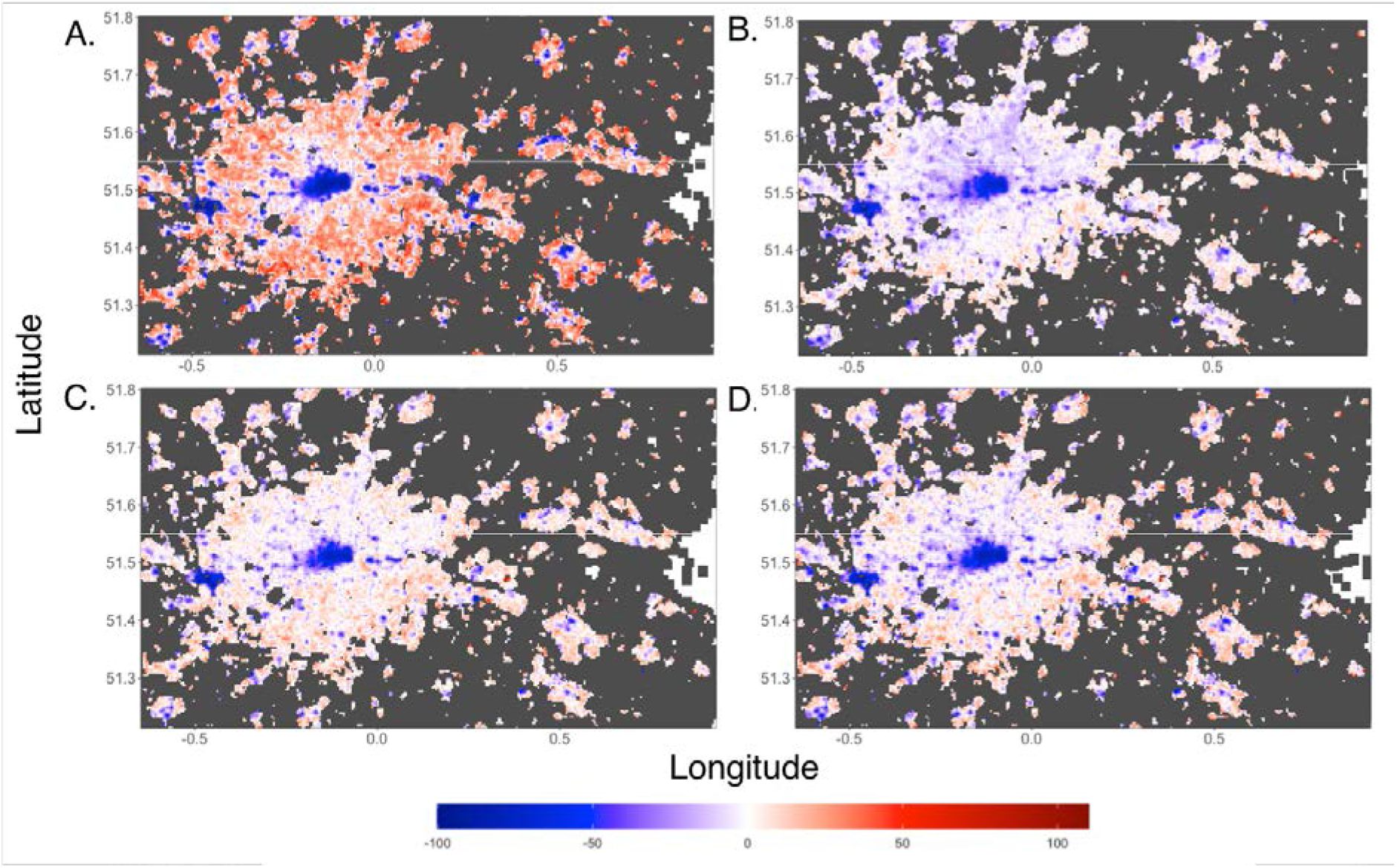
Relative changes in the average population density of daytime Facebook users within London under different mobility restrictions. A) Lockdown one (05/04/2020 - 12/05/2020). B) Summer 2020 (05/07/2020 - 31/08/2020). C) Lockdown two (05/11/2020 - 01/12/2020). D) Lockdown three (05/01/2021 - 08/03/2021). Time period is between 08:00 - 16:00 UTC. Data does not coincide with the beginning of lockdown one as data collection did not commence until 05/04/2020. The change is relative to a pre-COVID baseline (see Methods for details).

Regional population density for all regions excluding London generally increased or remained similar to the pre-COVID baseline during the first lockdown (Supplementary figure 1). However, increases in population density were not observed during lockdown two and three (Supplementary figure 1) where regional population density was generally lower than the pre-COVID baseline. London however showed large decreases in population density during lockdown one, and remained consistently lower than other regions (Supplementary figure 1). Within London, there were notable differences in the tile densities of Facebook users. During the first lockdown, central London showed large decreases, of the order of 80-90% in the population of Facebook users relative to the pre-COVID baseline period. In the more residential areas outside central London, increases in the relative number of Facebook users were observed (Figure 4A). During summer 2020 and throughout lockdown two and three the relative population density of Facebook users in central London remained lower than the pre-COVID baseline, however the increase in areas outside of central London was not as large.

The pandemic and mitigation measures extended beyond local restrictions to international travel as well, with a set of recommendations and restrictions implemented by the UK and destination countries. Figure 5 shows the major effects that these domestic and international travel guidelines and restrictions had on movements of Facebook users into and out of the UK. Changing restrictions across the study period lead to major fluctuations in travel patterns, with some countries showing substantial increases in traveller numbers followed by rapid decreases as restrictions were imposed.

**Figure 5.**
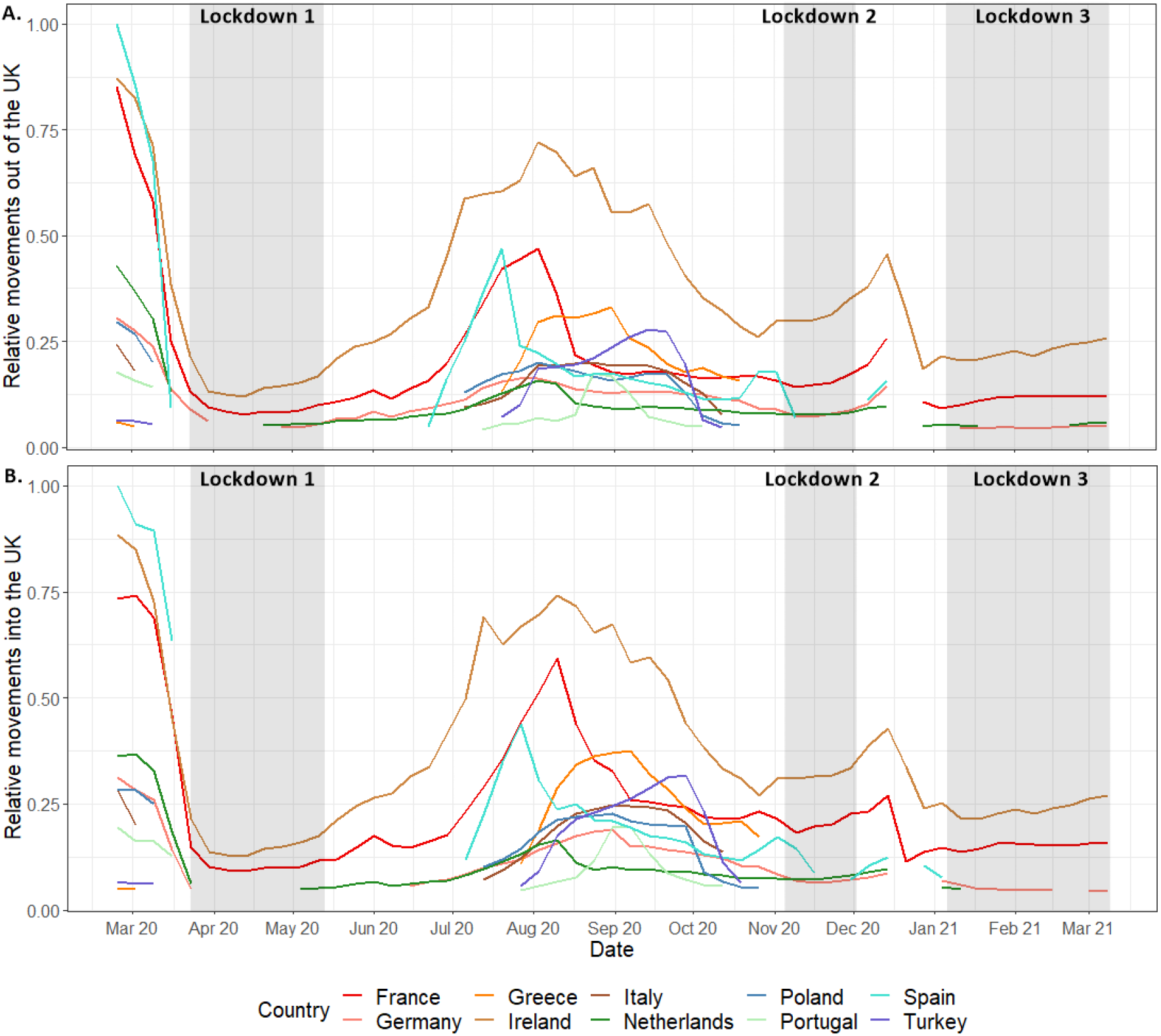
Weekly relative movement of Facebook users travelling (A) out of, and (B) into the UK for the 10 countries with the largest travel volumes across the study period. Gaps in the data represent periods where less than 1000 Facebook users travelled between the UK and another country. Movement is relative to the highest weekly movement in and out of the UK observed during the study period.

Across the first lockdown in March 2020, travel to and from all observed countries was greatly reduced. Following a relaxation of restrictions across the UK summer period (June - August 2020), travel to common holiday destinations such as Spain, France and Greece increased. In late July and August 2020, an increase in travel back into the UK from Spain, France and Greece can be observed from the top panel of Figure 5, potentially indicating holiday-makers returning in a hurry before restrictions are implemented. However, relative travel to and from many countries such as Germany and the Netherlands remained low across the study period. Over the study period, travel to and from Ireland was consistently higher than the other countries.

## Discussion

The COVID-19 pandemic has altered population mobility patterns globally since March 2020, and has likely resulted in some permanent shifts in commuting and international travel. Understanding these will be important for designing future pandemic preparedness and mitigation strategies, and the past year has highlighted the wide range of digital data sources that exist to aid this (Apple, 2020, Facebook, 2020, Google, 2020). Here we have explored what one of those data sources can tell us about changes that occurred across the UK in response to the pandemic and the policies implemented to mitigate its impacts.

The analyses presented here reveal heterogeneities across time and space in terms of changes to the travel patterns of Facebook users over the March 2020 - 2021 period, as well as some of the heterogeneities that exist within the UK in terms of population geography and behaviours. Compared to the 90 day period prior to 10th March 2020, it is clear that travel over relatively long distances between regions dropped substantially, particularly in the week prior to the implementation of the first lockdown, and during the first lockdown where travel flows were as much as 80% lower. This was not matched by movements over shorter distances, which, while reduced from normal levels, became more dominant as people remained within their local areas. Flows, distances travelled and co-location probabilities remained remarkably stable within each of the three ‘lockdown’ periods defined here, though clear increases in mobility patterns are seen in March 2021. The impact that each lockdown had on differences relative to the baseline period however were very different, with the second period resulting in only around 20-30% reduction in travel flows and distances generally compared to those of 60-70% in the first period. Figure 3 is revealing though, with each lockdown period exhibiting a more marked drop in co-location, and thus showing the desired impact of such interventions in reducing contacts. Co-location probabilities showed a peak for many regions over the Christmas period, and was likely a contributing factor to the growth of the major wave of infections seen from January 2021,

Local and regional differences were evident throughout the analyses. Though partially a function of where borders between regions were drawn, London remained an outlier throughout, with the largest reductions in travel flows and distances, but smallest reductions in co-location probabilities. Figure 4 highlights the extremes experienced in the region, with central London showing consistently lower densities throughout as workers, shoppers and tourists stayed away. In contrast, outer London showed substantially higher daytime densities, as working from home and local travel took over. While little evidence was seen for differing mobility patterns in regions of England under local restrictions, the other nations of the UK where types and timings of restrictions often differed to a degree, exhibited some strong variations compared to English regions. Scotland and in particular Northern Ireland maintained lower co-location probabilities throughout much of the study period, suggesting that the restrictions put in place in these nations potentially were more successful in reducing contact rates. Internationally, figure 5 paints a picture of unprecedented fluctuations over short time periods, as the UK government and those overseas wrestled with balancing the reopening of international tourism and business with the clear risks of infection importation (du Plessis et al., 2020; Lemey et al., 2021; Ruktanonchai et al., 2020).

A major source of uncertainty in translating the findings of the analyses presented here to population-wide inferences is the limited data on potential biases. While Facebook usage, smartphone ownership and use of location-enabled services has grown over the past few years, privacy restrictions mean that biases remain challenging to quantify, with the data potentially skewed towards younger and middle-aged groups, and not capturing the movements of children or the elderly. While there are likely biases in the data analysed that prevent conclusive inferences on population level insights, recent analyses have highlighted strong correlations with demographic variables from census-based data (Gibbs et al., 2021), giving confidence in its representativeness within the UK. Moreover, the patterns observed match those seen in alternative data sources (Jeffrey et al., 2020), and an ongoing area of future work is focussed on measuring these biases and extending dataset inter-comparisons. This will include the integration and comparison of data from call detail records with smartphone location history (e.g. Ruktanonchai et al, 2020), as well as examining associations with mobility data derived from surveys, census and other more traditional sources (e.g. Ruktanonchai et al, 2018; Wesolowski et al., 2013).

Another source of uncertainty arises from the lack of matching 2019 comparison data. UK population mobility patterns change significantly throughout a typical year (e.g. ONS 2018), and therefore, comparisons to a 90-day baseline period prior to March 10th 2020 do not provide a full picture of the extent of deviations from ‘normal’ patterns at each time of year. Moreover, changing Facebook usage throughout the study period may have impacted findings. The lockdown periods when people were often confined to their homes lead to a greater use of the internet, and social media in particular (Fritz & Kauermann, 2020, Mellon & Prosser, 2017). This may have led to different segments of the population being recorded in the data more often during different periods, and more movements captured in the lockdown periods. The differential privacy algorithms implemented by Facebook (Facebook, 2020) meant that varying levels of detail could be captured, which was a function of population and Facebook user densities. For example, Figure 4 showing detailed changes for London was not possible to produce for much of the UK where population densities were low, and subjective decisions were required on the scales of analyses undertaken and presented. Finally, the regional insights are subject to the modifiable areal unit problem (Openshaw 1983), whereby results are likely to be sensitive to the boundaries used.

The COVID-19 pandemic continues to be an unprecedented event in terms of numbers of people impacted around the World and changes to lifestyles. It is also unprecedented in terms of our abilities to observe and quantify many of those impacts through new ‘big’ digital datasets. Here we have shown what can be obtained through one of these data sources. While the data from Facebook are imperfect, valuable insights into how UK populations reacted to different interventions have been uncovered. These emphasise the potential of such sources to guide strategies in near-real time and support future pandemic planning.

## Data Availability

The data used in this study are proprietary data owned by Facebook. The data are not publicly available but are available to researchers by application to the Facebook Data for Good Partner Program (https://dataforgood.fb.com/).

## Acknowledgements

We would like to thank Alex Pompe and Laura McGorman at Facebook for their support with data access, processing, connections to end users and responding to questions. Additionally, we would like to acknowledge the COVID-19 Mobility Network (https://www.covid19mobility.org/) for support with data access and connections to data providers and policy makers. We also thank the team members at the UK Office for National Statistics Data Science Campus for providing guidance on insights from the data and discussing approaches to data processing. Finally, we thank the WorldPop group at the University of Southampton, in particular those in the project management and operations team for their support across this work.

## Author contributions

AJT conceived the analyses and lead the work undertaken. HERS, FSA, HMTC and AL undertook the download, management, processing and analysis of the data. All authors contributed to the design of the study and writing of the manuscript.

## Notes

### Competing Interest Statement

The authors have declared no competing interest.

### Funding Statement

No external funding was received for this work.

### Author Declarations

University of Southampton Ethics and Research Governance

### Summary of Updates

Updated references

