## Supporting information for "Domestic and international mobility trends in the United Kingdom during the COVID-19 pandemic: An analysis of Facebook data"

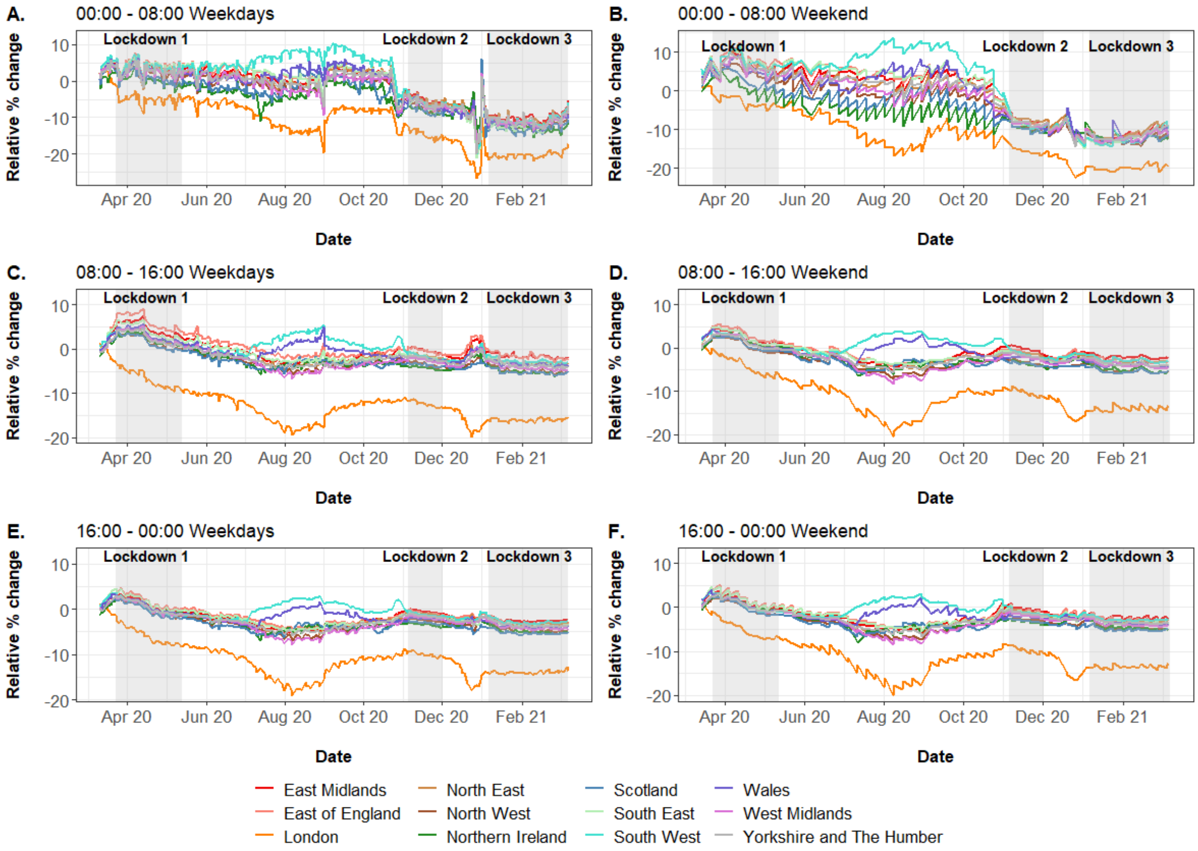


**Supplementary figure 1.** Percent change in population density of Facebook users within UK regions throughout 8-hour time periods on weekdays (A, C, E) and weekends (B, D, F) from March 10th 2020 and March 9th 2021. Regions are UK NUTS level 1 (Figure 1c). The change is relative to a pre-COVID baseline (see Methods for details). Shaded areas represent periods of national lockdown.

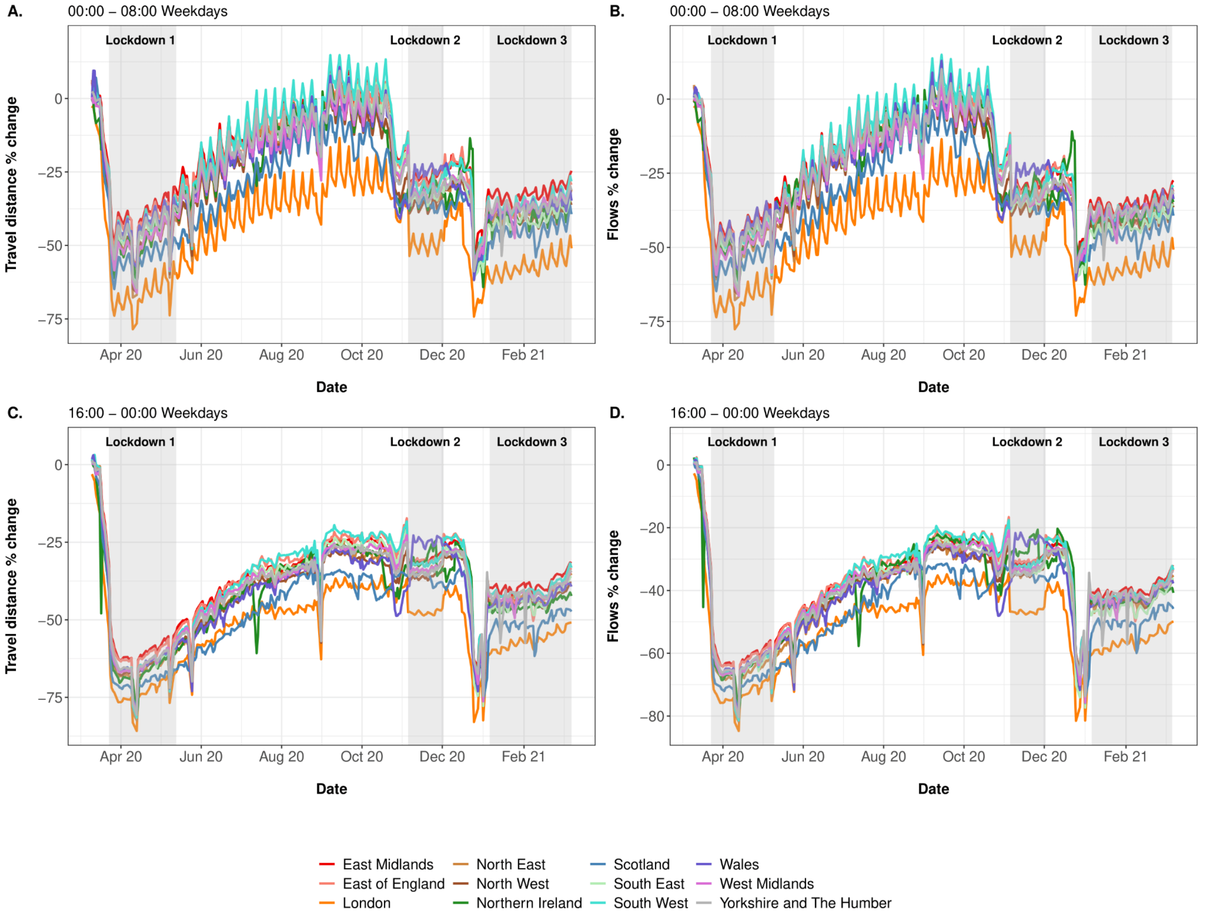


**Supplementary figure 2**. Percentage changes in (A, C) total distances travelled and (B, D) population movement (flows) for journeys between two adjacent 8-hour time periods on weekdays from March 10th 2020 to March 9th 2021. Regions are UK NUTS level 1 (Figure 1c). Time shown indicates the second 8-hour time period, where journeys ended. The change is relative to a pre-COVID baseline (see Methods for details). Shaded areas represent periods of national lockdown.

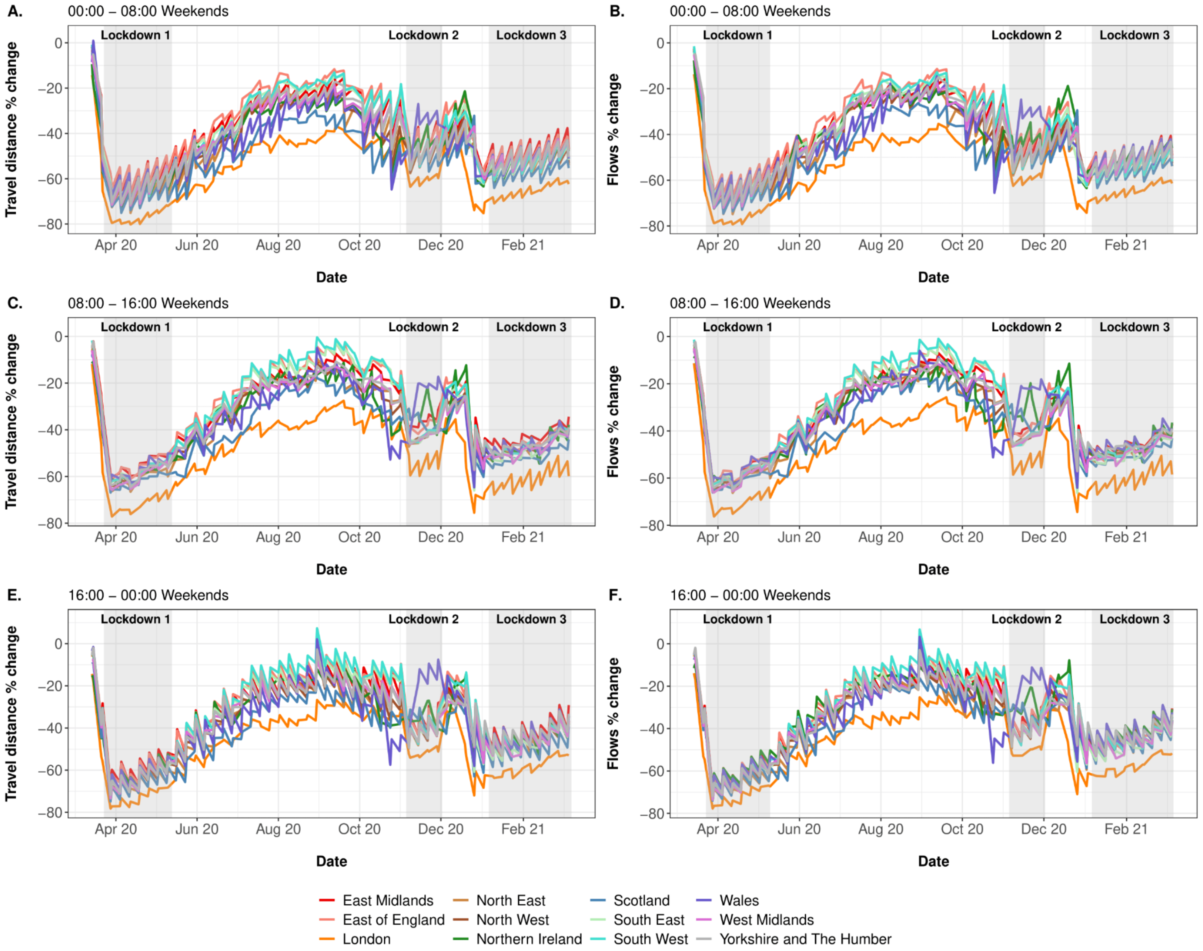


**Supplementary figure 3.** Percentage changes in (A, C, E) total distances travelled and (B, D, F) population movement (flows) for journeys between two adjacent 8-hour time periods on weekends from March 10th 2020 to March 9th 2021. Regions are UK NUTS level 1 (Figure 1c). Time shown indicates the second 8-hour time period, where journeys ended. The change is relative to a pre-COVID baseline (see Methods for details). Shaded areas represent periods of national lockdown.

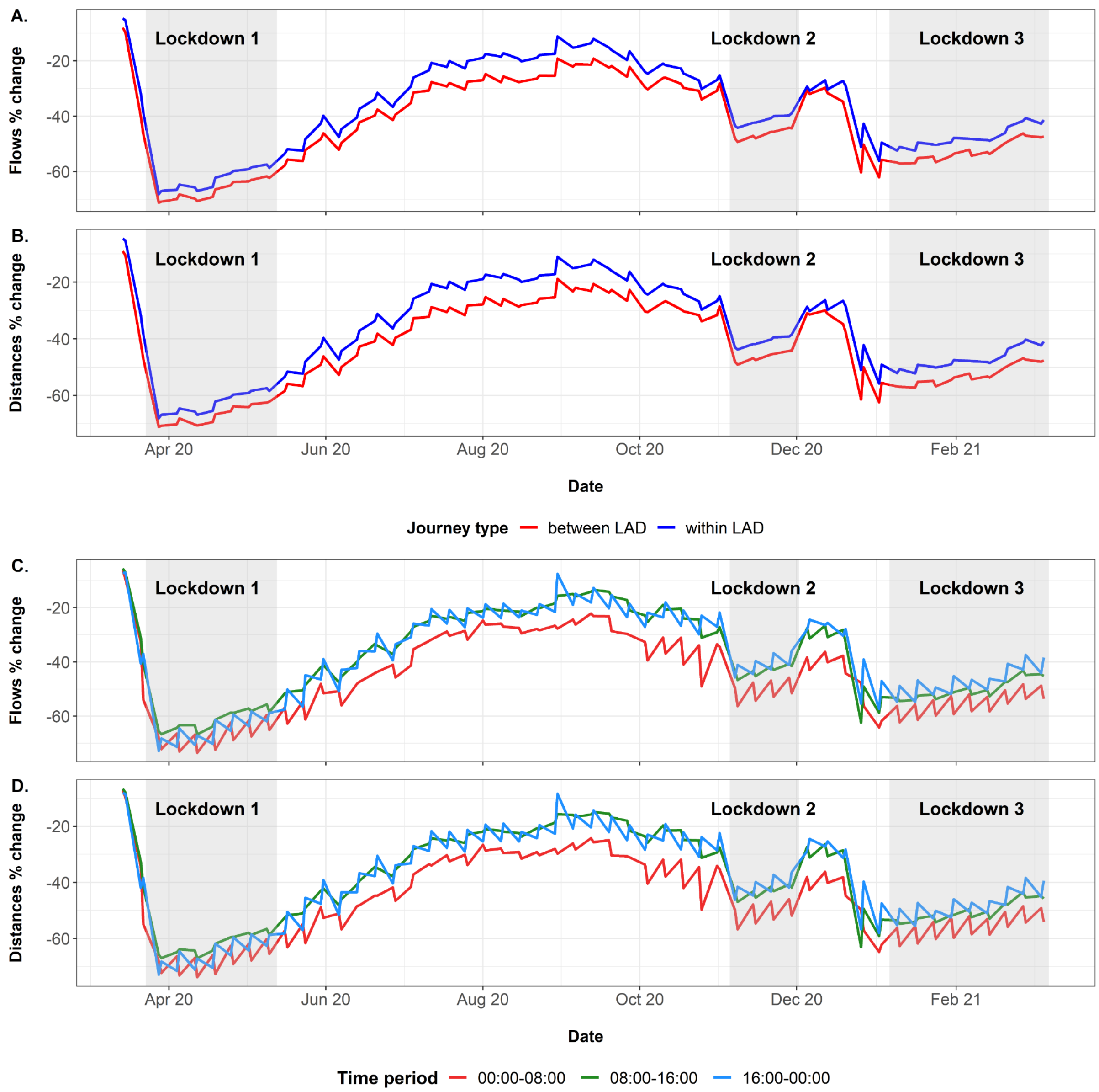


**Supplementary figure 4***.* Percentage changes in (A, C) daily movement (flows) of Facebook users and (B, D) total distance travelled from March 10th 2020 to March 9th 2021 on weekends. A and B show the changes corresponding to different journey types (between and within UK local authority districts). C and D show the changes corresponding to journeys ending within the different 8-hour time periods. The change is relative to a pre-COVID baseline (see Methods for details). Shaded areas represent periods of national lockdown.

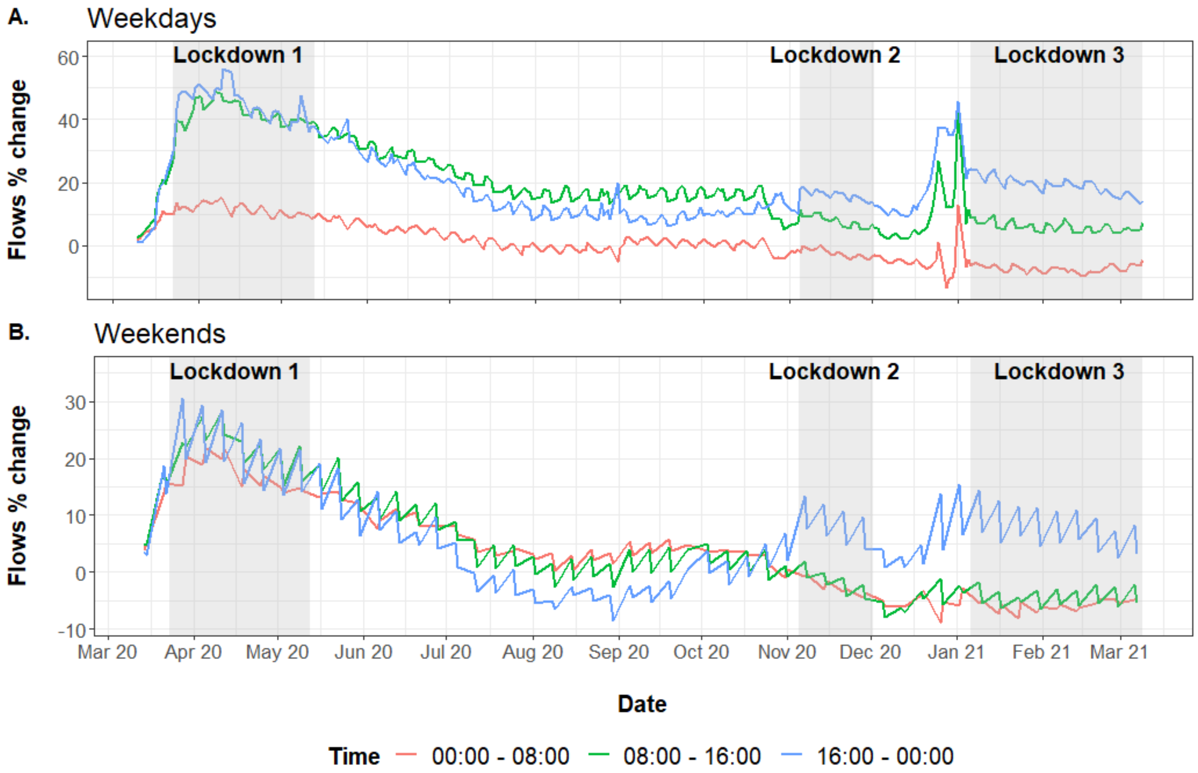


**Supplementary figure 5.** Percentage changes in daily number of Facebook users that remained within the same Bing tile locations between two adjacent 8-hour time periods from March 10th 2020 to March 9th 2021. The change is relative to a pre-COVID baseline (see Methods for details). Time shown indicates the second 8-hour time period. Shaded areas represent periods of national lockdown.

**Supplementary table 1.** Summary statistics of weekly trend of mean log co-location probabilities for the NUTS level 1 regions in the UK from March 2020 to March 2021.

| | Min. (date) | Q1 | Med. | Mean | Q3 | Max. (date) |
|---|---|---|---|---|---|---|
| **East Midlands (England)** | -18.193 (14/04/2020) | -17.131 | -16.859 | -16.602 | -15.985 | -14.869 (10/03/2020) |
| **East of England** | -18.891 (14/04/2020) | -17.595 | -17.358 | -17.083 | -16.464 | -15.128 (10/03/2020) |
| **London** | -18.366 (14/04/2020) | -16.8 | -16.327 | -16.228 | -15.507 | -14.13 (03/03/2020) |
| **North East (England)** | -18.947 (14/04/2020) | -17.837 | -17.567 | -17.241 | -16.544 | -15.152 (10/03/2020) |
| **North West (England)** | -18.32 (14/04/2020) | -17.188 | -16.898 | -16.571 | -15.818 | -14.455 (10/03/2020) |
| **Northern Ireland** | -19.905 (31/03/2020) | -18.945 | -18.632 | -18.54 | -18.361 | -15.962 (03/03/2020) |
| **Scotland** | -19.076 (31/03/2020) | -18.23 | -17.945 | -17.702 | -16.948 | -15.423 (10/03/2020) |
| **South East (England)** | -18.691 (14/04/2020) | -17.584 | -17.304 | -17.023 | -16.396 | -14.996 (10/03/2020) |
| **South West (England)** | -18.718 (14/04/2020) | -17.618 | -17.25 | -17.019 | -16.256 | -15.044 (10/03/2020) |
| **Wales** | -18.446 (14/04/2020) | -17.696 | -17.391 | -17.151 | -16.589 | -15.219 (10/03/2020) |
| **West Midlands (England)** | -18.084 (14/04/2020) | -16.985 | -16.685 | -16.454 | -15.796 | -14.618 (10/03/2020) |
| **Yorkshire and The Humber** | -18.507 (14/04/2020) | -17.391 | -17.061 | -16.779 | -16.05 | -14.87 (10/03/2020) |
